# Physical Health of Autistic Girls and Women: A Scoping Review

**DOI:** 10.1101/2020.05.16.20104216

**Authors:** Caroline Kassee, Stephanie Babinski, Ami Tint, Yona Lunsky, Hilary Brown, Stephanie H. Ameis, Peter Szatmari, Meng-Chuan Lai, Gillian Einstein

## Abstract

**Background:** There is a growing recognition of sex and gender influences in autism. Increasingly, studies include comparisons between sexes or genders, but few have focused on clarifying the characteristics of autistic girls’ and women’s physical health.

**Methods:** A scoping review was conducted to determine what is currently known about the physical health of autistic girls and women. We screened 1,112 unique articles, with 40 studies meeting the inclusion criteria. We used a convergent iterative process to synthesize this content into broad thematic areas.

**Results:** Overall, autistic girls and women experience more physical health challenges compared to non-autistic girls and women, and to autistic boys and men. Preliminary evidence suggests increased neurological conditions (e.g., epilepsy) in autistic girls and women compared to autistic boys and men. As well, the literature suggests increased endocrine/reproductive conditions in autistic girls and women compared to non-autistic girls and women.

**Limitations:** The literature has substantial heterogeneity in how physical health conditions were assessed and reported. Further, our explicit focus on physical health may have constrained the ability to examine potential interactions between mental and physical health. In addition, the widely differing research aims and methodologies make it difficult to reach definitive conclusions. Nevertheless, in keeping with the goals of a scoping review, we were able to identify key themes to guide future research.

**Conclusions:** Emerging themes in the literature suggest that autistic girls and women have heightened rates of physical health challenges compared to autistic boys and men, and non-autistic girls and women. Clinicians should seek to provide holistic care for this population that includes a focus on physical health and recognizes that autistic girls and women have co-occurring conditions that differ from those of autistic boys and men.

## Background

Autism spectrum disorder (hereafter autism) is a neurodevelopmental condition characterized by early-onset social-communication difficulties and repetitive, stereotyped behaviours. The estimated prevalence rate of autism is approximately 1% worldwide [1], more prevalent in males than females [2,3]. The widely reported male-to-female ratio for autism prevalence is 4-5:1, but large-scale population-based epidemiological studies suggest that the ratio is in fact lower at 3-4:1 [4], reflecting sex and gender differences in the likelihood of developing autism, which may be further accentuated by biases in clinical assessment and diagnoses [5].

Autism is highly associated with co-occurring health conditions [6]. It is hypothesized that this likely reflects complex epigenetic and pleiotropic gene-environment interactions and behavioural mechanisms [3,7], which are important to understand because they complicate the clinical presentation of autism. Co-occurring conditions are also associated with varied developmental trajectories [8,9] and unique social and psychological challenges that an individual experiences over the course of their lifetime [3]. Given the growing recognition of autism in girls and women, and that sex and gender differences in autism are important to study [10], it is critical to better understand the characteristics of co-occurring conditions in autistic girls and women.

Most research on co-occurring conditions in autistic girls/women relative to boys/men has focused on psychiatric conditions, suggesting increased internalizing psychopathology in autistic girls/women than in boys/men [11–13]. The latest meta-analysis on co-occurring psychiatric diagnoses in autistic people also shows that studies with a higher proportion of girls/women tend to find higher rates of depression [14]. However, less attention has been paid to sex and gender differences in autism outside the domain of mental health, especially regarding physical health^1^. Accurate and in-depth information in this domain, especially concerning autistic girls/women, is essential to the provision of comprehensive and sex- and gender-sensitive health care, and important for elucidating clinically useful sub-groups within the autism spectrum. In view of this, we conducted a scoping review of the literature, focused on the extent and range of research pertaining to physical health in autistic girls/women. Our research questions guiding the review were twofold: (1) *what do we know about the physical health of autistic girls/women;* and (2) *how specific are these physical health concerns to autistic girls/women, as compared to autistic boys/men, as well as non-autistic girls/women?*

## Methods

We conducted a scoping review of the literature following the methodological framework outlined by Arksey and O’Malley [15] and recent Preferred Reporting Items for Systematic Reviews and Meta-Analyses (PRISMA) standards for scoping reviews [16]. Scoping reviews allow a broad survey of the literature in a particular area, to determine existing themes and areas of inquiry that are under-researched. Scoping reviews typically do not conduct an assessment of bias in the research or with appraising or generating effect sizes [15]. We considered a scoping review to be the most appropriate approach for examining emerging evidence concerning the physical health of autistic girls/women, since it was unclear what specific questions should be posed in this area given the limitation of current literature. Therefore, our purposes were to summarize the extent and range of research pertaining to physical health in autistic girls/women and to identify evidence gaps. In this way, we surveyed all of the literature with respect to physical health in autistic girls/women, without exclusions based on specific samples or comparisons being made in the literature. Furthermore, as this literature often—unfortunately—conflates gender and sex, it is difficult to tease apart their respective effects. Hence, in interpreting the findings, references to “girls/women” were assumed to refer to biological (cis-gender) females and references to “gender” were read very carefully to determine whether they referred to biological sex, gender identity, or socially determined norms.

We systematically searched the following databases according to PRISMA standards [17]: CINAHL, PubMed, EMBASE, PsycINFO, Scopus, and Web of Science (see Appendix 1: Search Strategy). As this was a scoping review aimed at assessing general themes in the published literature, rather than analyzing specific types of data, gray literature was not included in the searches. Autism and co-occurring physical health conditions were defined using a combination of keywords and controlled vocabulary applicable to each database (see Appendix 1: Search Strategy). We purposely kept the definition of “physical health” as broad as possible, in order to gather a wide range of studies and gain a thorough coverage of the published literature with respect to non-mental health related conditions. There was no publication type or date restriction at this stage, but the search results were limited to human studies and journal articles written in English. The final database search was performed on December 5^th^, 2019 and references were managed using Mendeley (https://www.mendeley.com/).

A systematic selection process was used to determine the final articles included in this review. After duplicates were removed, two authors (CK and SB) screened titles and abstracts with support from senior authors (M-CL and GE), using broad criteria to allow for the inclusion of any potentially relevant study for further evaluation. Full-text articles were evaluated for inclusion by CK and SB. The pool of studies identified based on screening titles, abstracts, and consultations with senior authors determined the inclusion and exclusion criteria. At this stage, articles were included if they: (1) reported on co-occurring physical health conditions in people with a diagnosis of autism as defined by the DSM-IV, DSM-5 or ICD-10 criteria, or had direct relevance to physical health of autistic girls/women; (2) included a clearly articulated sex-specific or gender-specific description or analysis of these conditions; (3) studied biological females only, or if the total female autism sample size was ≥15 and with at least one-eighth (12.5%) of the total autism sample being biologically female (while seemingly arbitrary, these criteria ensured that included studies had a sufficient number of girls/women to derive sex-specific or gender-specific information); (4) reported original, English-language research articles or reviews published in peer-reviewed scientific journals; and (5) in the case of review articles, used systematic search methods and included sex-specific or gender-specific analyses and interpretation. Exclusion criteria included: (1) review articles using non-systematic search methodology; (2) opinion pieces; (3) editorials; (4) case reports; or (5) conference papers. Final decisions on which articles to include were made via discussion within the research team. Articles were grouped by main topic area and study design for organizational clarity. Data were extracted as shown in Tables 1 and 2, with relevant findings summarized in Results. We used a convergent iterative process involving multi-stage revisions among all authors to synthesize included studies into a series of thematic areas that broadly summarize the literature.

**Table 1.** Overview of included studies (n=40)

| **Year of Publication Range** | **Country of Origin** | **Main Topic of Study** | **Study Design** |
| --- | --- | --- | --- |
| 2007-2019 | North America = 16<br>Europe = 13<br>Asia = 5<br>Middle East = 3<br>Australia = 3<br>Africa = 0<br>South America = 0 | Prevalence rates for overall physical health conditions = 18<br>Neurological conditions = 4<br>Female-specific endocrine or reproductive conditions = 7<br>Gastrointestinal, metabolic, nutritional conditions = 6<br>Immunological conditions, immune factors = 3<br>Physical health review = 2 | Systematic reviews/meta-analyses = 5<br>Reviews with systematic search methods = 2<br>Cross-sectional studies, with population/registry samples = 23<br>Cross-sectional studies, with clinical/community samples = 10 |

**Table 2.** Summary of included studies (n=40, in alphabetical order)

| **Author Year**<br>**Country** | **Study Design** | **Topic Area** | **Main Focus** | **Sample Size** | **Age**<br>**(autism**<br>**sample)** | **% Female**<br>**(autism sample**<br>**by sex)** | **% ID**<br>**(autism**<br>**sample)** | **Comparison**<br>**Groups (against**<br>**autistic females)** |
| --- | --- | --- | --- | --- | --- | --- | --- | --- |
| Amiet et al.<br>2008 [35]<br>France | Systematic<br>review and<br>meta-meta-analysis | Neurological<br>condition –<br>epilepsy | Relative risk for<br>epilepsy in autism,<br>and associations<br>with ID | N=1,530 with<br>autism in<br>sex/gender<br>analyses from<br>14 studies | All ages | 22.2% in<br>sex/gender-<br>related analyses<br>(1,191 males,<br>339 females) | Not reported | Autistic males |
| Bitsika &<br>Sharples 2018<br>[43]<br>Australia | Cross-sectional<br>community<br>sample | Female-specific<br>endocrine<br>conditions | Effects of menarche<br>on sensory features<br>of autism | N=53 with<br>autism | 6 to 17<br>years | 100% (53<br>females) | Not reported | None |
| Ben-Itzhak et<br>al. 2013 [41]<br>Israel | Cross-sectional<br>clinic/communit<br>y sample | Neurological<br>conditions –<br>phenotypes | Sex differences in<br>neurological<br>phenotypes in<br>autism | N=663 with<br>autism | 1 to 15<br>years | 13.0% (577<br>males, 86<br>females) | 35.0%* | Autistic males |
| Bowers et al.<br>2015 [42]<br>USA | Cross-sectional<br>clinic/<br>community<br>sample | Neurological<br>conditions –<br>phenotypes | Phenotypes in<br>autism in preterm<br>vs. term births | N=883 with<br>autism | 0 to 18<br>years | 17.4% (728<br>males, 155<br>females) | N=853 with<br>data, 34.5% | Autistic males |
| Broder-Fingert et al.<br>2014 [52]<br>USA | Cross-sectional<br>registry sample | Gastrointestinal,<br>metabolic,<br>nutrition –<br>obesity | Overweight and<br>obesity prevalence<br>in autism | N=2,359 with<br>autism, N=3,696<br>controls | 2 to 20<br>years | 20.7% (2,359<br>males, 617<br>females) | Not reported | Autistic males |
| Cashin et al.<br>2018 [26]<br>Australia | Review with<br>systematic<br>methodology | Scoping review | Physical health in<br>adults with autism | N=3,882 with<br>autism, from 6<br>studies | 18+ years | Not reported | Not reported | Autistic males |
| Cawthorpe et<br>al. 2017 [32]<br>Canada | Cross-sectional<br>registry sample | Prevalence rates | Medical<br>comorbidities in<br>autism | N=2,040,<br>compared to<br>N=763,499 in<br>registry | All ages | 28.6% (1,457<br>males, 583<br>females) | Not reported | Same-sex general<br>population<br>controls |
| Cherskov et<br>al. 2018 [39]<br>USA | Cross-sectional<br>registry sample | Female-specific<br>endocrine<br>conditions –<br>PCOS | Polycystic ovary<br>syndrome (PCOS) | N=971 with<br>autism,<br>compared to<br>N=4,855 | Mean age<br>30.30<br>years (SD<br>9.1) | 100% | Not reported | General<br>population<br>women |
|  |  |  |  | controls |  |  |  |  |
| Chiang et al. 2015 [48]<br>Taiwan | Cross-sectional registry sample | Female-specific endocrine conditions – ovarian cancer | Risk of cancer in autistic people | N=8,438 with autism, N=76,332 controls | All ages | 17.9% (6,931 males, 1,507 females) | Not reported | General population women |
| Croen et al. 2015 [33]<br>USA | Cross-sectional registry sample | Prevalence rates | Medical and psychiatric conditions in autism | N=1,507 with autism, compared to N=15,070 general population controls | Adults (mean age 29.0 years (SD 12.2)) | 26.9% (1,102 males, 405 females) | 19.2% | Same-sex general population controls |
| Davignon et al. 2018 [29]<br>USA | Cross-sectional registry sample | Prevalence rates | Medical and psychiatric conditions in youth | N=4,123 with autism, N=20,615 with ADHD, N=2,156 with diabetes, N=20,615 general population controls | 14 to 25 years | 19.3% (3,326 males, 797 females) | 13% | Autistic males and females |
| Ewen et al. 2019 [36]<br>USA | Cross-sectional registry sample | Prevalence rates | Neurological conditions – epilepsy | N=6,975 with autism, in two cohorts (N=4,801 and N=1,736) | 6 to 18 years | 18.7% (5,671 males, 1,304 females) | 20.8% | Autistic males |
| Fortuna et al. 2016 [23]<br>USA | Cross-sectional clinical sample | Prevalence rates | Health conditions | N=255 with autism; no control | 18 to 71 years | 24.7% (192 males, 63 females) | N=141 with data; 91% with ID | Autistic males |
| Garcia-Paster et al. 2018 [54]<br>USA | Cross-sectional clinical sample | Gastrointestinal, metabolic, nutritional conditions | Obesity and physical activity in autistic children and adults | N=78 with autism | 7 to 48 years | 28.2% (56 males, 22 females) | Not reported | Autistic males, autistic children and adults |
| Guo et al. 2019 [50]<br>China | Cross-sectional clinical sample | Gastrointestinal, metabolic, nutritional conditions | Vitamin A and D deficiencies in autism | N=332 with autism, N=197 controls | Mean 4.87 years (SD 1.53) | 13.8% (286 males, 46 females) | Not reported | Autistic males |
| Hamilton et al. 2011 [45] | Cross-sectional community | Prevalence rates | Female-specific endocrine | N=124 with autism | 10 to 25 years | 100% | Not reported | None |
| USA | sample |  | conditions –<br>menstruation |  |  |  |  |  |
| Hand et al.,<br>2019 [34] | Cross-sectional<br>registry sample | Prevalence rates | Physical health in<br>older adults with<br>autism | N=4,685 with<br>autism,<br>N=46,850<br>controls | 65 years<br>and older | 32.3% (3,175<br>males, 1,510<br>females) | 43.8% | Same-sex general<br>population<br>controls |
| USA |  |  |  |  |  |  |  |  |
| Hu et al. 2019<br>[56] | Cross-sectional<br>registry sample | Immunological<br>conditions | Plasma levels of<br>cytokines in autism | N=87 with<br>autism, N=41<br>controls | 1 to 6<br>years | 17.2% (72<br>males, 15<br>females) | Not reported | Autistic males |
| China |  |  |  |  |  |  |  |  |
| Ingudomnukul<br>et al. 2007<br>[38] | Cross-sectional<br>clinic/communit<br>y sample | Female-specific<br>endocrine<br>conditions | Testosterone-related medical<br>conditions in autism | N=54 with<br>autism, N=74<br>mothers of<br>autistic girls,<br>and N=183<br>general<br>population<br>women<br>(controls) | 19 to 63<br>years | 100% | Not reported | Same-sex general<br>population<br>controls |
| UK |  |  |  |  |  |  |  |  |
| Jones et al.<br>2016 [24] | Cross sectional<br>clinical sample | Prevalence rates | Medical conditions<br>in autism | N=92 with<br>autism, no<br>controls | 23.5 to<br>50.5 years | 25% (69 males,<br>23 females) | N=82 with<br>data; 70%<br>with ID | Autistic males |
| USA |  |  |  |  |  |  |  |  |
| Masi et al.<br>2017 [55] | Cross-sectional<br>clinical sample | Immunological<br>conditions<br>and/or factors | Cytokine levels and<br>severity of autism<br>and other clinical<br>traits | N=144 with<br>autism | 2 to 18<br>years | 21.5% (113<br>males, 31<br>females) | Not reported | Autistic males |
| Australia |  |  |  |  |  |  |  |  |
| Mason et al.<br>2018 [22] | Cross-sectional<br>registry sample | Prevalence rates | Predictors of<br>quality of life in<br>autism | N=370 with<br>autism, no<br>controls | 17 to 80<br>years | 42.7% (199<br>males, 158<br>females, 13 not<br>reported) | Not reported | Autistic males |
| UK |  |  |  |  |  |  |  |  |
| Memari et al.<br>2012 [28] | Cross-sectional<br>community<br>sample | Prevalence rates | Comorbidities in<br>school children | N=91 with<br>autism | 6 to 14<br>years | 25% (68 males,<br>23 females) | Not reported | Autistic males |
| Iran |  |  |  |  |  |  |  |  |
| Moseley et al.<br>2018 [26] | Systematic<br>review and<br>meta-analysis | Prevalence rates | Sex differences in<br>autistic symptoms | N=254 with<br>autism, N=707<br>general<br>population<br>controls | Weighted<br>mean<br>36.37<br>years<br>(females<br>37.4 (SD<br>14), males | 53.5% (118<br>males, 136<br>females) | 0% | Autistic males |
| UK |  |  |  |  |  |  |  |  |
|  |  |  |  |  | 35.3 (SD 13.4) |  |  |  |
| Mouridsen et al. 2013 [40]<br>Denmark | Cross-sectional registry sample | Prevalence rates | Neurological conditions – cerebral palsy | N=4,180 with Asperger’s syndrome | 4 to 31 years | 17.9% (3,431 males, 749 females) | 0% | Autistic males |
| Pohl et al. 2014 [39]<br>UK | Cross-sectional registry sample | Female-specific endocrine conditions | Sex-linked steroid conditions in autism | N=415 with autism, N=415 age-matched controls | Mean age 36.39 years (SD 11.98) | 100% | Not reported | General population women |
| Rossignol & Frye 2012 [51]<br>USA | Systematic review and meta-analysis | Gastrointestinal, metabolic, nutritional conditions | Mitochondrial disease (MD) in autism | N=112 with autism-MD; N=536 autism only, from 65 studies | 0 to 20 years | 39% in autism-MD; 19% in autism only (no individual breakdown) | Not reported | Autism and autism-MD groups |
| Rubenstein et al. 2015 [31]<br>USA | Review with systematic methodology | Review article | Sex differences in co-occurring conditions in autism | Ranges N=28 to N=337,000, from 69 studies | Not reported | Not reported | Not reported | Autistic males |
| Rubenstein et al. 2018 [25]<br>USA | Cross-sectional registry sample | Prevalence rates | Temporal trends in co-occurring disorders in autism | N=6,379 with autism | 8 year-olds | 18.0% (5,230 males, 1,149 females) | 4.3% | Autistic males |
| Rydzewska et al. 2019a [20]<br>UK | Cross-sectional registry sample | Prevalence rates | Prevalence rates for comorbid categories | N=25,063 autism, N=1,523,756 controls | 0 to 24 years | 20.7% (19,880 males, 5,183 females) | 14.9% | Autistic males |
| Rydzewska et al. 2018 [19]<br>UK | Cross-sectional registry sample | Prevalence rates | Prevalence rates for comorbid categories | N=6,649 with autism; N=3,746,584 controls | 25+ years | 30.7% (4,610 males, 2,039 females) | 29.4% | Autistic males |
| Rydzewska et al. 2019b [18]<br>UK | Cross-sectional registry sample | Prevalence rates | General health status in autism | N=6,649 with autism, N=3,746,584 controls | 25+ years | 30.7% (4,610 males, 2,039 females) | 29.4% | Autistic males |
| Saghazadeh et al., 2019 [57]<br>Iran | Systematic review and meta-analysis | Immunological conditions and/or factors | Pro-inflammatory cytokines in autism | 38 studies with N=1,393 with autism, and N=1,094 controls | All ages | 36 out of 38 studies with sex/gender information; 1,584 males and 446 females total reported (autism and control); % of autistic females not reported | Not reported | Autistic males, general population controls |
| Stacy et al. 2014 [21]<br>USA | Cross-sectional registry sample | Prevalence rates | Differences in co-occurring diagnoses in males and females with autism | N=913 with autism | Weighted mean age 9.91 years (females 10.82 (SD 0.61) vs. males 9.71 (SD 0.27)) | 18.3% (746 males, 167 females) | Not reported | Autistic males |
| Steward et al. 2018 [44]<br>USA | Cross-sectional community sample | Female-specific endocrine conditions – menstruation | Autistic women’s experiences with menstruation | N=123 with autism, N=114 controls | 16 to 60+ years | 100% | Not reported | General population women |
| Sundelin et al. 2018 [47]<br>Sweden | Cross-sectional registry sample | Female-specific endocrine conditions | Pregnancy in autistic women | N=1,382 women with autism (N=2,198 births), N=503,486 general population controls (N=977,742 births) | Adults of child-bearing ages | 100% | Not reported | General population women |
| Supekar et al. 2017 [27]<br>USA | Cross-sectional registry sample | Prevalence rates | Sex differences on comorbid conditions in autism | N=4,790 with autism | All ages | Not reported for overall sample | Not reported | Autistic males |
| Tseng et al. 2018 [49]<br>Taiwan | Systematic review and meta-analysis | Gastrointestinal, metabolic, nutritional conditions | Iron deficiency and autism | N=1,603 with autism across 3 analyses, N=1,507 controls in meta-analyses, from 25 studies | 0 to 18 years | 20.0% (individual breakdown not reported) | Not reported | Autistic males |
| Viscidi et al. 2013 [37] | Cross-sectional registry sample | Prevalence rates | Neurological conditions – epilepsy | N=5,815 with autism | All ages, majority between 4- | 17.5% (4,800 males, 1,015 females) | 15.5% | Autistic males |
| USA |  |  |  |  | 12 years (~75%) |  |  |  |
| Yang et al. 2018 [53]<br>China | Cross-sectional clinic/community sample | Gastrointestinal, metabolic, nutritional conditions | GI symptoms in autism, association with sleep | N=169 with autism, N=172 controls | Mean 5.23 years (SD 2.0) | 14.2% (145 males, 24 females) | Not reported | Autistic males |

\*Calculated using mean and standard deviations from IQ scores, for males and females
SD = standard deviation
SIR = standardized incidence ratio
OR = odds ratio
RR = relative risk ratio
CI = confidence interval
GI = gastrointestinal

## Results

### Search Results

We screened a total of 1,112 unique citations, and reviewed the full-text of 201 articles, with 40 studies ultimately meeting the inclusion criteria (Figure 1). The majority of the studies were from North America and Europe, cross-sectional, and about general prevalence rates for health conditions in autism (Table 1). The papers studied autistic individuals of all ages and functional levels (Table 2), with comparisons of autistic girls/women to (1) autistic boys/men, (2) non-autistic girls/women, (3) girls/women with other neurodevelopmental or psychiatric conditions, or (4) descriptive studies of autistic girls/women only (Table 2).

**Figure 1.**
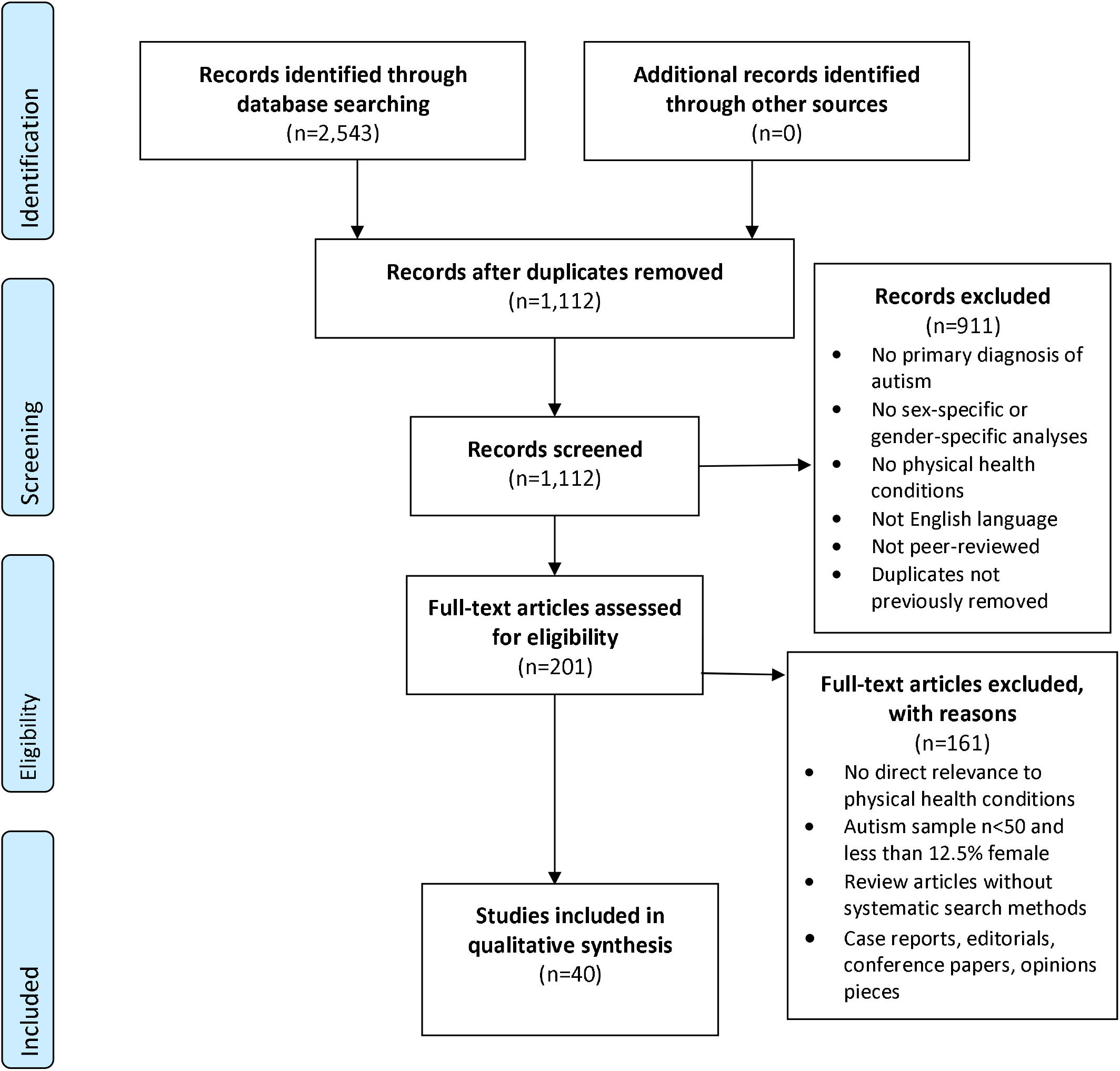
PRISMA Flow Diagram for Study Selection.

We identified five key themes with mixed findings emerging. Among these, three had relatively more consistent findings, including (1) *Autistic girls/women may experience more overall physical health conditions than autistic boys/men and non-autistic girls/women;* (2) *Neurological conditions, especially epilepsy, are more prevalent in autistic girls/women compared to autistic boys/men;* and (3) *Autistic girls/women may experience more menstruation-related complications, and female-specific endocrine and reproductive conditions, compared to non-autistic girls/women*. The rest two themes showed (4) *inconsistent evidence for gastrointestinal and related conditions in autistic girls/women as compared to autistic boys/men*, and (5) *possible evidence for differences in immune profiles for autistic girls/women compared to autistic boys/men*.

### Summary of Key Themes

#### 1. Autistic girls/women may experience more overall physical health conditions than autistic boys/men and non-autistic girls/women

##### a. Comparing autistic girls/women to autistic boys/men

The majority of studies described prevalence rates for a wide range of physical health conditions in autistic people (Table 1). Out of these, several studies reported prevalence of physical health conditions in autism by sex, comparing autistic girls/women directly to autistic boys/men [18-21]. Autistic girls/women have higher odds than autistic boys/men for six disorder categories derived from census data, including three relevant to physical health: deafness, blindness, and physical disability [19,20]. These results were reported in two population-based studies focusing on n=25,063 autistic children and youth aged 0-24 years (Odds Ratio, OR autistic girls/women compared to autistic boys/men: deafness 2.07 [95% CI 2.04-2.10], blindness 2.51 [2.12-2.97], and physical disability 2.60 [2.50-2.71]) [20]; and for n=6,649 autistic adults (deafness 1.169 [95% CI 1.001-1.365], blindness 1.232 [1.051-1.443], and physical disability 1.504 [1.333-1.697]) [19]. In a follow-up study [18] on the same adult cohort in [19], census questions on general health were used to identify physical health conditions among autistic adults. The study found that among young adults (25-34 years), autistic women were more likely to have poorer health compared to autistic men (43.9% autistic women vs. 35.7% autistic men reporting “poor general health”; χ^2^ =13.2, df=1, p<0.001) [18]. This men-women difference was not statistically significant in other age ranges. Supporting this, a study of 913 autistic children (weighted mean age 9.9 years) by Stacey et al. from a registry sample on nine conditions, two about physical health (epilepsy and hearing problems), found no differences between autistic girls and boys [21].

Other studies examined the associations between physical health indicators and sex/gender [22–25]. Mason et al. analysed sex (which was referred to as gender) as a predictor of quality of life in n=370 autistic adults (17-80 years) from a research registry cohort, finding statistically significant interactions between gender and the physical subscale of the World Health Organization Quality of Life assessment, indicating lower physical-related quality of life in autistic women (*M*=45.98 [*SD*=19.57]) than in autistic men (*M*=52.98 [*SD*=17.32], p=0.019) [22]. Fortuna et al. surveyed overall health and functional status in n=255 autistic adults (18-71 years) and found that female sex (referred to as gender) was associated with lower odds of good or excellent overall health (OR for autistic women 0.5 [95% CI 0.2-1.0] with autistic men as a reference group) [23]. Jones et al. found increased medical comorbidity associated with female sex (referred to as gender) (autistic women median 16 conditions vs. 10 for autistic men, p=0.01) in n=92 autistic adults (23-50 years) in a community sample [24]. In contrast, Rubenstein et al. examined co-occurring conditions in n=6,379 eight year-old autistic children across eight US treatment sites over 2002-2010 to estimate the percentage of autistic children with co-occurring conditions in four broad categories, including neurological conditions [25]. Rates of change for neurological conditions in autistic children over 2002-2010 were the same for autistic boys and autistic girls.

Studies comparing autistic girls/women to autistic boys/men also examined physical health conditions and symptoms to quantify differences [26–29]. Moseley et al. conducted a meta-analysis including n=254 autistic adults (weighted mean age 36.4 years) to determine sex differences in self-reported autistic characteristics, including sensorimotor symptoms (some of which related to physical health, such as sensitivity to pain) [26]. Results revealed significantly more severe sensorimotor symptoms in autistic women than in autistic men (t[252]=4.346, p<0.001). Supekar et al. compared prevalence rates in n=4,790 autistic people and n=1,842,575 non-autistic individuals of all ages from large medical registry cohorts, for a range of co-occurring conditions including physical health, such as epilepsy, inflammatory bowel disease, bowel disorder, and muscular dystrophy [27]. They found higher prevalence for epilepsy in autistic girls/women (18.54%) than in autistic boys/men (15.14%, p<0.05); however, this finding was modulated by age, such that epilepsy was female-predominant in 0-18 years and 18-35 years, but male-predominant in >35 years of age. Also, while bowel disorders exhibited higher male prevalence in autism overall, there was statistically significant higher female prevalence in >35 years of age (23.08% female vs. 10.00% males, p<0.05). In a small community-based sample of n=91 children (6-14 years), Memari et al. found that autistic girls had higher prevalence of neurological conditions than autistic boys (~46% in girls vs. ~19% [extracted from graph, exact numbers not given in the report] in boys, p=0.02) [28]. Davignon et al. described the prevalence of co-occurring conditions in n=4,123 autistic youth (14-25 years) from a California-based clinical registry, noting that most physical health conditions were more common in autistic girls/women than in autistic boys/men, although no statistical comparison was done [29].

Finally, two reviews surveyed physical health in autistic girls/women compared to autistic boys/men [30,31]. Cashin et al. conducted a scoping review on the physical health status of n=3,896 autistic adults from 6 relevant studies, noting only 3 included sex-specific analyses, with inconsistent results [30]. Rubenstein et al. reviewed sex differences in developmental, medical and psychiatric conditions for autistic people of all ages, with samples ranging from n=28 to n=337,000 from 69 studies, with 20 studies reporting specifically on the physical health domain [31]. They outlined insufficient research (hence evidence) on sex differences in autism for most conditions.

##### b. Comparing autistic girls/women to non-autistic girls/women

Three studies computed ORs for autistic girls/women relative to non-autistic girls/women, and contrasted these to ORs for autistic boys/men relative to non-autistic boys/men in the general population [32-34]. Cawthorpe et al. [32] and Croen et al. [33] reported ORs for autistic females and males compared to same-sex general population controls, using the International Classification of Diseases Ninth Edition (ICD-9) diagnostic codes for medical disorders, in registry-based autism samples (all age ranges) of n=2,040 autistic individuals (referencing to n=763,499 general population controls) [32] and n=1,507 autistic adults (referencing to n=15,070 general population controls) [33]. Both studies found that autistic girls/women had increased odds for most conditions compared to non-autistic girls/women [32,33]. There were also potential sex-differential patterns in the physical conditions that were elevated compared to same-sex non-autistic individuals. For example, complications during mothers’ pregnancy/childbirth (autistic male OR 1.52 [95% CI 1.07-2.15], autistic female OR 0.55 [0.44-0.68]) and genitourinary system diseases (autistic male 1.2 [1.08-1.33], autistic female 0.99 [0.81-1.20]) [32], autoimmune diseases (autistic male 1.30 [1.01-1.68)], autistic female 1.12 [0.78-1.60]) and gastrointestinal disorders (autistic male 1.50 [1.25-1.79], autistic female 1.05 [0.80-1.39]) [33] were only elevated in autistic boys/men. In contrast, other conditions were elevated only in autistic girls/women, including blood and blood-forming organ disorders (autistic female OR 1.35 [1.11-1.65], autistic male OR 1.14 [0.96-1.35]) and endocrine, nutritional, metabolic diseases and immunity disorders (autistic female 1.47 [1.25-1.73], autistic male 0.63 [0.56-0.71]) [32], and stroke (autistic female 4.97 [1.46-16.86], autistic male 1.48 [0.59-3.70]) [33]. Finally, Hand et al. examined prevalence of physical health conditions in n=4,685 older autistic adults (≥65 years) enrolled in US-based Medicare, compared to n=46,850 general population controls [34]. They found increased odds for most conditions for autistic women compared to same-sex general population controls, and there were no conditions elevated only in one sex but not the other. The three physical health conditions with the largest ORs in autistic women were epilepsy (20.8 [17.7-24.4]), Parkinson’s disease (8.2 [6.2-10.7]), and other gastrointestinal conditions (4.6 [4.1-5.1]).

#### 2. Neurological conditions, especially epilepsy, tend to be more prevalent in autistic girls/women compared to autistic boys/men

In addition to studies reporting on general health condition prevalence rates including epilepsy mentioned in Theme 1 [27,34], several studies focused specifically on neurological conditions reported a higher prevalence of epilepsy in autistic girls/women compared to autistic boys/men [35–37]. Amiet et al. conducted a systematic review and meta-analysis on sex differences in epilepsy and autism, including 14 studies with n=1,530 autistic individuals (across all ages) to assess the pooled risk ratio (RR) for epilepsy by sex, with findings showing lower rates of epilepsy in autistic boys compared to autistic girls (RR 0.55 [95% CI 0.45-0.66], p<0.001; 34.5% in autistic girls/women vs. 18.5% in autistic boys/men) [35]. Two other studies reporting medical conditions in autism found elevated rates of epilepsy in autistic women compared to non-autistic women. Ingudomnukul et al. reported epilepsy prevalence at 7.4% in n=54 autistic women (mean age 38.2 years) vs. 1.1% in n=183 age-matched non-autistic women (p<0.05) [38]; Pohl et al. found 4.1% epilepsy prevalence in n=415 autistic women (mean age 36.37 years) vs. 1.4% in n=415 age-matched non-autistic women (p=0.016) [39].

Two studies further characterized epilepsy in autism [36,37]. Ewen et al. explored associations between epilepsy and autism severity in two cohorts from a US-based research registry, totalling n=6,975 autistic children (6-18 years) [36]. They found a higher risk for epilepsy in autistic girls compared to autistic boys in one cohort, with n=4,801 autistic individuals (RR 1.32 [1.14-1.52], p<0.05) but no statistically significant findings in the other, smaller cohort (n=1,736 autistic individuals). They also found independent positive associations between epilepsy and severity indicators such as intellectual disability, language impairment, core autism symptom, and motor dysfunction. Viscidi et al. explored the links between autism and epilepsy by examining prevalence and clinical characteristics of n=5,815 autistic individuals (all ages, ~75% of the sample between 4-12 years) from four research registry cohorts [37]. In addition to finding that epilepsy co-occurring with autism was associated with older age, lower cognitive ability, poor adaptive language functioning, developmental regression, and more severe autism symptoms, they found that epilepsy was more prevalent in autistic girls/women than in autistic boys/men (7.0% in autistic girls/women vs. 3.9% in autistic boys/men, p<0.001).

Another three studies characterized other neurological conditions with respect to sex [40–42]. Mouridsen et al. examined prevalence of cerebral palsy in a nation-wide cohort of n=4,180 autistic individuals with ICD-10 Asperger’s syndrome (4-31 years) [40]. They found increased cerebral palsy in Asperger’s syndrome (0.65%) than in the general population (0.17%), but no significant difference between autistic girls/women and boys/men (0.80% in autistic girls/women vs. 0.61% in autistic boys/men, p=0.56). Ben-Itzchak et al. studied specific neurologic phenotypes in autism in n=663 autistic children (1-15 years) from community samples, and reported no sex differences in autism severity, cognitive ability or adaptive functioning [41]. Neurological anomalies were more prevalent in autistic girls than in autistic boys, including microcephaly (15.1% vs. 4.5%, χ^2^ =15.0, df=1, p<0.001) and minor neurological-musculoskeletal deficits (73.8% vs. 57.1%, χ^2^=8.0, df=1, p<0.001), but no significant sex differences were found for seizures or macrocephaly. Finally, Bowers et al. characterized the phenotypes of n=883 autistic children (0-18 years) born preterm and at term, to examine comorbidities including seizure disorders; these were more frequent among autistic boys born preterm vs. those born term (17.0% vs. 8.5%, p=0.01), whereas no such preterm-term differences were found in autistic girls [42].

#### 3. Autistic girls/women may experience more menstruation-related complications, and female-specific endocrine and reproductive conditions, compared to non-autistic girls/women

Eight studies examined female-specific endocrine or reproductive conditions in autism [38,39,43–48]. Three studies specifically examined menstruation in women with autism [43–45]. Two of these administered web-based surveys to understand menstruation experiences of autistic girls/women [45] and compare these experiences with those of non-autistic women [43,44]. They identified menstruation-related complications and notable differences in menstruation experiences between autistic and non-autistic girls/women. Hamilton et al. surveyed n=124 autistic girls/women (10-25 years) online by parent-report and self-report questionnaires, and found that girls/women commonly experienced symptoms of dysmenorrhea (91%) and pre-menstrual syndrome (96%), and 33% endorsed autism-associated difficulties during the menstrual cycle (increased irritability/aggression before menses), worsening of autistic behaviours, and increased repetitive movements and obsessive behaviours [45]. Steward et al. surveyed n=123 autistic women (16-60+ years) and compared their responses to n=114 age-matched non-autistic women using qualitative synthesis of written survey responses [44]. Although there were many overlaps in menstrual cycle issues between autistic and non-autistic women, autistic women highlighted autism-specific issues, including a cyclical amplification of autism-related challenges, sensory differences and emotional regulation challenges, which had significant negative impact on their lives. Bitsika and Sharpley investigated the effects of menarche on the sensory features of autism, in n=53 autistic girls (6-17 years), using clinical questionnaires completed by their mothers [43]. Autistic girls who had reached menarche had lower sensation seeking (less sensory interests) (F(_25_,_27_)=2.113, p=0.030) and multisensory processing (F(_7_,_45_)=3.187, p=0.008) compared to those who had not yet reached menarche.

Additionally, two studies focused on female-specific endocrine and reproductive health conditions. There was a higher prevalence of irregular menstruation (57.4% vs. 28.6%, p<0.001) and painful periods (44.4% vs. 28.0%, p<0.05) in autistic women compared to non-autistic women (n=54 autistic women, n=183 non-autistic women) [38]. Unusually painful periods (39.3% vs. 26.3%, p=0.00004) and pre-menstrual syndrome (in contraceptive pill users) (24.0% vs. 13.8%, p=0.001) were also more common in autistic women (n=415 autistic women, n=415 non-autistic women) [39].

Along with menstruation cycle complications, five studies explored female-specific endocrine conditions or physical health conditions in relation to reproductive health in autism, and reported a range of autism-specific findings [38,39,46–48]. Ingudomnukul et al. reported on testosterone-related conditions in n=54 autistic women (19-63 years), compared to n=74 mothers of autistic children, and n=183 age-matched, non-autistic women using a self-reported clinical questionnaire administered online [38]. Autistic women, compared to non-autistic women, had higher rates of polycystic ovary syndrome (PCOS, 11.3% vs. 2.7%, p<0.05), delayed puberty (7.4% vs. 0.5%, p<0.01), and hirsutism (29.6% vs. 4.4%, p<0.001). Similarly, using a self-reported clinical questionnaire, Pohl et al. examined reproductive and sexuality-related characteristics and conditions in n=415 autistic women (mean age 36.37 years) compared to n=415 age-matched non-autistic women [39]. Based on response patterns, two groups (named by the authors as “typical” and “steroidopathic”) were identified, with the prevalence of the “steroidopathic group” significantly increased in the autism compared to control groups (∆G =15, df=1, p=0.0001). In particular, there were higher frequencies of reproductive and steroid-linked conditions in autistic women, including irregular menstrual cycle (46.3% vs. 34.0%, p=0.0002), severe acne in non-contraceptive pill users (21.3% vs. 5.9%, p=0.002), precocious puberty (3.1% vs. 0.5%, p=0.003), and early growth spurt (20.2% vs. 12.8%, p=0.002) [39]. Cherskov et al. conducted three interrelated studies on the associations between autism and PCOS using nation-wide electronic health records. One examined the risk for PCOS in n=971 autistic women (mean age 30.3 years) vs. n=4,855 non-autistic women [46]. The prevalence of PCOS in autistic women was higher than in non-autistic women (2.3% vs. 1.1%, p<0.01; OR 2.01 [1.22-3.30]). Sundelin et al. reported on pregnancy outcomes in n=2,198 births to n=1,382 autistic women of child-bearing age, compared to n=877,742 births in n=503,846 women from the general population using data from a nation-wide registry [47]. They found autistic women were at increased risk for pre-eclampsia (OR 1.34 [1.08-1.66]); for outcomes of pregnancy and the fetus, autistic women were at increased risk of giving birth preterm (OR 1.30 [1.10-1.54]), medically indicated preterm birth (OR 1.41 [1.08-1.82]), and elective caesarean delivery (OR 1.44 [1.25-1.66]). Finally, Chiang et al. investigated risk of cancer in n=8,438 autistic children, youth and young adults (0-25+ years) compared to n=76,332 general population controls, from nation-wide registries [48]. They found higher standardized incidence ratios (SIR) for ovarian cancer in autistic women compared to the general population (SIR 9.21 [95% CI 1.12-33.29]).

#### 4. Inconsistent evidence for differences in gastrointestinal, metabolic, and nutritional conditions between autistic girls/women and autistic boys/men

Six studies compared gastrointestinal, metabolic, nutritional and related conditions between autistic girls/women and boys/men [49–54]. Yang et al. examined the prevalence of gastrointestinal problems in n=169 autistic children (mean age 5.23 years) and reported that autistic girls had greater likelihood of gastrointestinal problems than autistic boys (OR 3.88 [1.33-11.35], p=0.013); further, more gastrointestinal symptoms were correlated with more severe core autistic symptoms [53].

Two studies examined nutritional deficiencies in autism. In a systematic review and meta-analysis of peripheral iron levels and iron deficiency in autistic children (25 articles on peripheral ferritin, hair iron and, food iron intake in n=1,603 children (0-18 years)), Tseng et al. found no significant associations between sex and iron levels in the autism group [49]. Guo et al. assessed vitamin A and vitamin D deficiencies in n=332 autistic children (mean age 4.87 years), compared to n=197 age-matched controls from community samples [50]. They reported that autistic girls had significantly lower serum 25-OH vitamin D than autistic boys (p<0.05).

Rossignol and Frye performed a systematic review to determine the prevalence and characteristics of mitochondrial disease in autistic children, identifying 65 studies for qualitative synthesis, with 18 publications totalling n=536 autistic children and youth, and n=112 with autism and mitochondrial disease (0-20 years) [51]. Rates for specific clinical features, including female sex, were found to be elevated in the autism-mitochondrial disease group compared to the autism-only group (39% female in autism-mitochondrial disease group vs. 19% female in autism-only group, χ^2^= 18.7, p<0.0001).

Finally, two studies examined obesity. Broder-Fingert et al. compared n=2,976 autistic children and youth (2-20 years) to n=3,696 age-matched controls on sex- and age-adjusted body mass index (BMI), calculating odds for being overweight and obese compared to controls [52]. Autistic girls were less likely to be obese compared to autistic boys (OR 0.71 [0.55-0.93]), but this did not hold for overweight (OR 1.06 [0.81-1.39]). Garcia-Paster et al. compared obesity and physical activity status in n=44 autistic children and adolescents (7-18 years) and n=34 autistic adults (19-48 years), finding that overweight and obesity were significantly more prevalent in autistic men vs. autistic boys (p<0.001) and autistic men than in autistic women (p=0.035) [54].

#### 5. Possible differences in immune profiles for autistic girls/women compared to autistic boys/men

Three studies reported on sex-specific immunological conditions or immune factors in autism. Masi et al. measured the levels of 27 cytokines using multiplex assay in n=144 autistic children and adolescents (2-18 years) from a registry-based sample [55]. They reported that in autistic girls reduced levels of IL-1β, IL-8, MIP-1β, PDGF-BB and VEGF were associated with increased autism symptoms, while in autistic boys this was the case only of reduced PDGF-BB. The authors concluded that cytokine expression in autism was moderated by sex. Hu et al. measured plasma levels of 11 cytokines in n=87 autistic children (1-6 years), compared to n=41 age-matched non-autistic children, to characterize immune profiles and their association with autistic symptoms [56]. They found overall, autistic children had higher plasma levels of Eotaxin, TGF-β1, and TNF-α than non-autistic children. In autistic girls, only the increase in Eotaxin was statistically significant, whereas in autistic boys, the most consistent increase was in TGF-β1. Finally, Saghazadeh et al. conducted a meta-analysis on circulating concentrations of pro-inflammatory cytokines in n=1,393 autistic individuals (of all ages) compared to n=1,074 non-autistic controls, extracted from 38 studies [57]. Findings indicated higher concentrations of pro-inflammatory cytokines IFN-γ, IL-1β, IL-6, and TNF-α in autistic individuals than in controls; meta-regression revealed a correlation with sex as well as differences in mean serum levels of certain pro-inflammatory cytokines in autism, including IL-1β and TNF-α.

## Discussion

The purpose of this scoping review was to explore what is known about co-occurring physical health conditions in autistic girls and women. Out of the 201 articles reviewed full-text, only 40 met our inclusion criteria, mainly due to the paucity in reporting on sex or gender differences among populations with autism and the low percentages of autistic girls/women included in the current literature. This highlights a male-biased lens and relative ignorance of women’s health and female experiences in the scientific and clinical knowledge about physical health and autism so far. There is a pressing need for more research that includes large numbers of autistic girls/women in order to better understand their physical health. This should be prioritized in order to advance the best clinical care for autistic individuals [10].

Some emerging patterns of co-occurring physical health conditions are worth further examination and replication. With respect to Theme 1, the current literature suggests that autistic girls/women overall tend to have more physical health challenges and lower overall health and quality of life than do autistic boys/men [18–20,22,23,32–34]. However, with the exception of neurological conditions (especially epilepsy) it is still unclear which specific conditions are more prevalent in autistic girls/women compared to autistic boys/men or to non-autistic girls/women. Such inconsistency could be related to the substantial heterogeneity in the autism population even within each sex [58] or could be related to confounding factors (e.g., genetic mutation load or other neurodevelopmental disabilities). Based on Theme 2, epilepsy is the most studied condition, found to be more prevalent in autistic girls/women than in autistic boys/men in most included studies [23,27,28,31,33,35]. However, the complexity underlying this association remains, with studies highlighting potential confounding factors such as heightened autism symptom, language impairment, motor dysfunction, intellectual disability in autistic girls/women [36,37], or multifactorial etiology and shared neurological abnormalities [59]. On the other hand, barriers experienced by girls/women to receive an autism diagnosis [2,4,10] may result in autistic girls/women without evident developmental disabilities being under-represented in the current literature. Therefore, potential differences between autistic boys/men and autistic girls/women suggested by the current literature may be influenced by increased phenotypic complexity and severity among diagnosed autistic girls/women compared to autistic boys/men. It remains unclear if the same male-female differences hold in autistic individuals who are so far under-recognized and un-diagnosed.

There is some evidence indicating increased female-specific endocrine or reproductive health concerns in autistic girls/women (e.g., sensory, menstruation-related, hormonal conditions, and ovarian cancer), and potentially sex-specific immune profiles, as highlighted in Theme 3 and Theme 5. Nevertheless, these findings should be viewed as preliminary owing to the moderate sample sizes (Table 2) [43–45] and the reliance on self-report questionnaires to characterize female-specific endocrine conditions rather than direct clinical assessments [38,39,43–45]. Interestingly though, both themes can be hypothesis-generating and have implications for plausible biological mechanisms underlying autism, endocrine, and immune alternations to be investigated in future research.

For Theme 3, some have speculated that endocrine dysregulation in autistic girls/women is partly indicative of altered prenatal sex-steroid exposure [60,61], which may contribute to both endocrine dysregulation and autism-related neurodevelopmental and behavioural characteristics [62–64], with some emerging empirical support published recently [65–69]. How such prenatal endocrine factors contribute to the mechanisms leading to autism and concurrent physical health disorders in a sex-differential manner remains unclear, and is an area requiring more in-depth mechanistic investigation. Equally, there is growing evidence supporting the role of multidirectional interactions between prenatal immune activation, epigenetic regulation in key brain regions, and postnatal environments, in producing a range of distinct but related autistic phenotypes [70,71]. It is possible that there are shared mechanisms underlying autism and co-occurring endocrine and immune alterations, with sex-differential mechanisms involved.

As well, findings regarding gastrointestinal, metabolic, and nutritional conditions (Theme 4) require much more research to elucidate. There are preliminary indications that gastrointestinal [72] and metabolic/nutritional conditions [73], including obesity and diabetes [74], are of particular relevance to autistic people, especially girls/women. These conditions could involve shared etiological mechanisms with autism as well as with life experiences living with autism.

### Clinical Implications

The finding that autistic girls/women having more physical health challenges than non-autistic girls/women and autistic boys/men is of crucial clinical relevance. Improving physical health is integral to the care of all autistic individuals [75,76]. Frontline and primary care clinicians should regularly attend to and resolve unmet health care needs for autistic people, for children, youth and adults alike [77,78] and more specifically for autistic girls/women [79], particularly regarding (but not restricted to) neurological, endocrine/reproductive, immune and metabolic conditions. Incorporating existing knowledge on women’s health to the autism population is essential and will significantly enrich sex- and gender-informed health care for autistic people. Conversely, improved attention to physical health in girls/women who also experience difficulties in social-communication, restricted/stereotyped behaviours and sensory issues might facilitate the identification of later-recognized autism in girls/women [80]. As with other women and men, sex and gender strategies need to be applied to their health across the lifespan.

Another key consideration is the interplay between physical and mental health. Autistic people are prone to experience mental health challenges (which we did not review here) [14]. However, many psychiatric medications for such challenges have side effects that are more commonly experienced in autistic than non-autistic individuals [75,81–83] contributing to heightened risk to physical health (e.g., weight gain and endocrine problems related to psychotropic medications). These findings have not yet been sufficiently studied in a sex-specific manner. Meanwhile, physical health challenges (e.g., epilepsy, hormonal dysregulation) can have detrimental impact on mental health and affect mood and behaviour. Such complexity and interplay may result in the high clinical needs and multiple service use that are common in the autism population, particularly girls/women [84–86]. Many of these physical health challenges are treatable with the proviso that clinical trials need to disaggregate their data by sex, which is unfortunately still insufficiently done for clinical trials involving autistic people. Timely diagnosis and treatment will enhance wellbeing associated with both physical and mental health of autistic individuals across the lifespan. This review has revealed that autistic girls/women are a unique population with unique needs from autistic boys/men and non-autistic girls/women. Therefore, it behooves us to develop comprehensive services that integrate developmental, mental and physical health for autistic girls/women.

## Limitations

There are several limitations to our review. First, it is possible that we were unable to identify all studies relevant to our guiding question due to the heterogeneity in how physical health conditions are assessed and reported in the literature. Nevertheless, based on the principles of a scoping review, we have identified potential areas in the literature that warrant future investigation and areas with insufficient information as yet to make firm conclusions (as outlined earlier in Discussion). Second, the decision to focus on physical health thus excluding studies only focusing on psychiatric co-occurring conditions meant that we could not explore how mental and physical health are intertwined in autistic people, particularly in girls/women. Finally, our scoping review results demonstrate that our understanding in the physical health of autistic girls/women is still emerging. The limited number of studies in each thematic area, varying quality and research methodologies make it difficult to reach definitive conclusions.

However, several lessons can be learned from our review. There is a lack of consistent, basic epidemiological information on the prevalence and incidence for co-occurring health conditions in the autism population by sex and in particular by gender. Additionally, there is insufficient longitudinal studies to chart the emergence of co-occurring conditions, randomized control trials to address these conditions, and a lack of biological studies to elucidate mechanisms implicated in the development of physical health challenges in autism, by sex and by gender. Further, there is a significant under-representation of autistic girls/women in most studies, and only a small minority of studies formally examine and report sex/gender-differential effects in their primary analyses. Finally, it is extremely rare in the current empirical literature that sex (biological attributes) and gender (psychological, social and cultural attributes) are defined and examined separately in valid ways. These gaps need to be addressed in future research, alongside clarification of sources of demographic, clinical and etiological heterogeneity such as age, pubertal stage, developmental trajectories, intellectual functioning, and genetic background. Such clarification is fundamental for future studies to generate etiological and mechanistic insights by studying co-occurring physical conditions in autism, with sex- and gender-informed lenses.

## Conclusions

To our knowledge, this is the first scoping review on physical health in autistic girls/women. Emerging themes suggest that autistic girls/women may have heightened rates of co-occurring physical health challenges compared to autistic boys/men and non-autistic girls/women. Therefore, clinicians should provide holistic care that integrates not only developmental and mental health, but also physical health. Future studies need to include sufficient numbers of autistic girls/women to achieve adequate power, attend to physical health and the intertwined nature of developmental, mental and physical health, and use sex- and gender-informed lenses.

**List of Abbreviations**
PRISMA: Preferred Reporting Items for Systematic Reviews and Meta-Analyses ICD: International Classification of Diseases DSM: Diagnostic and Statistical Manual OR: Odds Ratio
SIR: Standardized Incidence Ratio
RR: Risk Ratio
SD: Standard Deviation

## Data Availability

The articles included in this scoping review are available from the corresponding author.

## Declarations

### Ethics approval and consent to participate

Not applicable

### Consent for publication

Not applicable

### Availability of data and materials

The articles included in this scoping review are available from the corresponding author.

### Authors’ contributions

M-CL, GE and SB conceived and planned the study. CK and SB carried out the literature search, screening, extraction and summary of data. M-CL and GE supervised the study and contributed to literature screening and summary of findings. CK, SB, M-CL and GE drafted the manuscript. AT, YL, HB, SHA and PS contributed to the interpretation of findings and writing of the manuscript. M-CL and SHA obtained funding support for the study. CK and SB contributed equally as first authors. M-CL and GE contributed equally as senior authors.

## Acknowledgements

Not applicable

## Footnotes

1 We define *physical health* in this context to encompass non-mental health conditions within the broad category of medical disorders or problems.

